# Effectiveness and waning of protection with different SARS-CoV-2 primary and booster vaccines during the Delta pandemic wave in 2021 in Hungary (HUN-VE 3 study)

**DOI:** 10.1101/2022.04.14.22273898

**Authors:** Zoltán Vokó, Zoltán Kiss, György Surján, Orsolya Surján, Zsófia Barcza, István Wittmann, Gergő Attila Molnár, Dávid Nagy, Veronika Müller, Krisztina Bogos, Péter Nagy, István Kenessey, András Wéber, Lőrinc Polivka, Mihály Pálosi, János Szlávik, György Rokszin, Cecília Müller, Zoltán Szekanecz, Miklós Kásler

## Abstract

**Background:** In late 2021, the pandemic wave was dominated by the Delta SARS-CoV-2 variant in Hungary. Booster vaccines were offered starting from August 2021.

**Methods:** The nationwide HUN-VE 3 study examined the effectiveness and durability of primary immunization and single booster vaccinations on SARS-CoV-2-related infection, hospitalization and mortality during the Delta wave.

**Results:** The study population included 8,087,988 individuals aged 18–100 years at the beginning of the pandemic. During the Delta wave, after adjusting for age, sex, calendar day, and chronic diseases, vaccine effectiveness (VE) of primary vaccination against registered SARS-CoV-2 infection was between 11% to 77% and 18% to 79% 14–120 days after primary immunization in the 16–64 and 65–100 years age cohort respectively, while it decreased to close to zero in the younger age group and around 40% or somewhat less in the elderly after 6 months for almost all vaccine types. In the population aged 65–100 years, we found high, 88.1%–92.5% adjusted effectiveness against Covid-19 infection after the Pfizer-BioNTech, and 92.2%–95.6% after the Moderna booster dose, while Sinopharm and Janssen booster doses provided 26.5%–75.3% and 72.9%–100.0% adjusted VE, respectively. Adjusted VE against Covid-19 related hospitalization was high within 14–120 days for Pfizer-BioNTech: 76.6%, Moderna: 83.8%, Sputnik-V: 78.3%, AstraZeneca: 73.8%, while modest for Sinopharm: 45.7% and Janssen: 26.4%. The waning of protection against Covid-19 related hospitalization was modest and booster vaccination with mRNA vaccines or the Janssen vaccine increased adjusted VE up to almost 100%, while the Sinopharm booster dose proved to be less effective. VE against Covid-19 related death after primary immunization was high or moderate: for Pfizer-BioNTech: 81.5%, Moderna: 93.2%, Sputnik-V: 100.0%, AstraZeneca: 84.8%, Sinopharm: 58.6%, Janssen: 53.3%). VE against this outcome also showed moderate decline over time, while booster vaccines restored effectiveness up to almost 100%, except for the Sinopharm booster.

**Conclusions:** The HUN-VE 3 study demonstrated waning VE with all vaccine types for all examined outcomes during the Delta wave and confirmed the outstanding benefit of booster vaccination with the mRNA or Janssen vaccines. This is the first study to provide comparable effectiveness results for six different booster types during the Delta pandemic wave.

## 1 Introduction

In Hungary, six different SARS-CoV-2 vaccines were approved in the first half of 2021, and five of them were investigated and showed high or very high short-term effectiveness against SARS-CoV-2 infection (Alpha variant) and Covid-19 related mortality in the nationwide HUN-VE 1 study (1). In the second half of 2021, a growing number of studies started reporting a waning effectiveness for vaccines over time, especially against the new, more infectious Delta variant and to less extent against Covid-19 related death (B.1.617.2) (2,3,4,5,6). To maintain protection against emerging new waves and variants, several European countries including Hungary started offering booster vaccine doses in summer 2021 (7,8). The benefit of booster vaccination has been demonstrated by a number of recent studies (9,10,11). On August 1, 2021, the Hungarian government introduced the option of a booster dose with one of six vaccine types at least 4 months after primary immunization, particularly for the vulnerable population such as people aged 60 years or older and those having chronic illnesses (12).

The aim of our study was to estimate the effectiveness of six different vaccine types as well as their combinations as primary or booster vaccines against SARS-CoV-2 infection, Covid-19 related hospitalization and death during the Delta pandemic wave between September 2021 and December 2021 in Hungary. We also aimed to evaluate the durability of protection offered by vaccine combinations to provide guidance for countries where multiple vaccine types are available.

## 2 Methods

The study population included Hungarian residents aged 18 to 100 years who were registered in the Hungarian-COVID-19 Registry based on the National Public Health Center (NPHC) and National Health Insurance Fund Manager (NHIFM) database on March 4, 2020, when the first case of Covid-19 infection was detected in Hungary. Exclusion criteria included any inconsistencies in data such as a person receiving two different vaccine types for primary immunization, missing information on the type of the second vaccine dose, first vaccination administered before the first potential date of administration, fewer than 14 days between the first and second doses, a date of diagnosis before the first case was officially reported, or a date of death preceding the date of first vaccination. Fewer than 1,000 cases were excluded for reasons other than age outside the predefined range.

Cases of SARS-CoV-2 infection were reported on a daily basis using a centralized system via the National Public Health Center (NPHC). The report is based on (i) Covid-19-related symptoms identified by hospital physicians and general practitioners, (ii) positive nucleic acid amplification test reported by microbiological laboratories. Cases identified by symptoms were confirmed by PCR or antigen test included in the EC rapid test list (13). Covid-19 related hospitalization was defined as hospitalization with a positive PCR or antigen test within 5 days before or 20 days after admission to hospital. Covid-19 related mortality was defined as death during SARS-CoV-2 positivity without previously declared recovery and without another clear cause of death (e.g., accident, suicide). The definition was based on WHO recommendations and defined by the healthcare government (14).

Individuals were classified as primary vaccinated if at least 14 days had passed since the administration of the second dose of the BNT162b2 (Pfizer-BioNTech), HB02 (Sinopharm), Gam-COVID-Vac (Sputnik-V), AZD1222 (AstraZeneca), or mRNA-1273 (Moderna) vaccines, or the first dose of Ad26.COV2.S (Janssen). At any time point during the study period, the unvaccinated, control population included individuals who had not received any dose of any Covid-19 vaccine type beforehand.

Vaccine effectiveness (VE) was defined as 1 minus the incidence rate ratio of the outcome in question. Vaccine combinations administered to at least 3,000 people by December 31, 2021 or with at least 300 cases from the start of the pandemic until December 31, 2021 in Hungary were included in the analysis. Vaccine combinations were further broken down into subcategories according to the time elapsed since their administration. These subcategories were defined as 14–120, 121–180, 181– 240, or more than 240 days. For booster vaccinations, only the 14–120 days category was established, because there were very few individuals during the Delta wave who had received their booster vaccination more than 120 days earlier.

All Hungarian residents aged 18–100 years who were registered in the NHIFM on March 4, 2020 were considered unvaccinated and uninfected persons apart from the first identified cases of Covid-19 infection. For each day afterwards until the end of the study period, we calculated the number of individuals at risk for different groups by age, sex, history of certain chronic diseases and vaccination, taking into account the types of vaccines and the time elapsed since their administration (intervention). Individuals who died were removed from the study base. The numbers of new registered infections, COVID-19 related hospitalizations, and deaths were registered for each day according to intervention, and individuals experiencing an outcome were removed from the study population of the respective analysis. Person-days for each intervention group were calculated by adding up the number of persons in each group for each day.

Age was categorized as 18–24, 25–34, 35–44, 45–54, 55–64, 65–74, 75–84, and 85–100 years. The presence of chronic diseases was identified based on in-and outpatient health service utilization and prescription data from the NHIFM between January 1, 2013 and March 3, 2021. The following chronic conditions were considered: cardiovascular diseases (myocardial infarction, angina, chronic heart failure, peripheral vascular disease, and stroke), diabetes mellitus (type 1 and type 2), immunosuppression (immunosuppressive therapy and transplantation), chronic pulmonary diseases (asthma and chronic obstructive pulmonary diseases), neoplasms, and chronic kidney diseases. The definitions of chronic diseases are presented in Supplementary Table 1.

Incidence rates (number of outcomes divided by person-time of observation) together with their exact 95% confidence intervals (CIs) were calculated for the period between September 13, 2021 and December 31, 2021 (Delta wave). Rate ratios with their exact confidence intervals were obtained considering unvaccinated as the reference category, using STATA (version 16.1).

Mixed effect negative binomial regression model was used to derive adjusted incidence rate ratios (IRRs) with 95% CIs for each outcome adjusted for age, sex, history of different chronic diseases, and calendar day (modelled as a random effect), which is better suited for over-dispersed count data than the traditional Poisson regression. The model is a random intercept model, which allows for different incidence rates in the reference category (i.e., unvaccinated) each day, but assumes fixed effect of the intervention categories. Separate models were fitted to estimate age group specific effects (i.e., 18–64 years and 65–100 years).

The study was approved by the Central Ethical Committee of Hungary (OGYÉI/10296-1/2022 and IV/1722-1/2022/EKU).

## 3 Results

The study population included 8,087,988 individuals (18-64 years: n=6,193,552, 65-100 years: n=1,894,436) at the beginning of the pandemic.

In the age group of 16–64 years, the crude incidence rate of SARS-CoV-2 infection was 67.6 per 100,000 person-days in the unvaccinated cohort during the Delta wave. In the primary vaccination cohorts, crude incidence rates varied between 5.2 and 38.3 per 100,000 person-days after 14–120 days of the second vaccine dose (Supplementary Table 2). Crude incidence rates of infection progressively increased 120–180, 181–240 and >240 days after the second vaccine dose for each vaccine type, even exceeding crude incidence rates of the unvaccinated population 6 months after primary vaccination. On the other hand, after booster vaccination with the Pfizer-BioNTech, Moderna or Janssen vaccines, crude incidence rates fell below 20 per 100,000 person-days, regardless of the type of vaccines used for primary immunization except for the Sputnik-V, Sputnik-V, Janssen combination. On the other hand, booster vaccination with the Sinopharm vaccine resulted in crude incidence rates of 25.2–61.2 per 100,000 person-days, depending on the type of the primary vaccination (Supplementary Table 2).

Similar results were seen in the 65–100 years age cohort: lower crude incidence rates of SARS-CoV-2 infection 14–120 days after primary vaccination for each vaccine type compared to the unvaccinated cohort (54.8 per 100,000 person-days), increasing infection rates after 4 months, and very low rates (<10 per 100,000 person-days) after booster vaccination with mRNA vaccines, regardless of primary vaccine types (Supplementary Table 3).

Higher effectiveness and slightly less waning of protection were found against Covid-19 related hospitalization and death in both age groups. However, both hospitalization and mortality rates were close to respective rates in the unvaccinated cohort 240 days after the second vaccine dose for most vaccine types, while booster vaccination led to relevant decreases in the incidence rates of these outcomes (Supplementary Tables 4–7).

After adjusting for age, sex, chronic diseases, and calendar day, vaccine effectiveness of primary vaccination against SARS-CoV-2 infection varied between 10.9% and 76.9% 14–120 days after the second dose in the 16–64 years age cohort (Figure 1A), and between 17.8% and 79.1% in the 65–100 years age cohort (Figure 2A). Adjusted vaccine effectiveness decreased to close to zero in the younger age group and around 40% or somewhat less in the elderly after 6 months for almost all vaccine types. In the population aged 65–100 years, we found high, 88.1%–92.5% adjusted effectiveness against Covid-19 infection after the Pfizer-BioNTech booster dose, and 92.2%–95.6% adjusted effectiveness after the Moderna booster during the first 4 months after booster immunization. The Sinopharm and Janssen boosters provided 26.5%–75.3% and 72.9%–100.00% adjusted effectiveness, respectively, depending on primary immunization type (Figure 1A and 2A, Supplementary Table 3).

**Figure 1.**
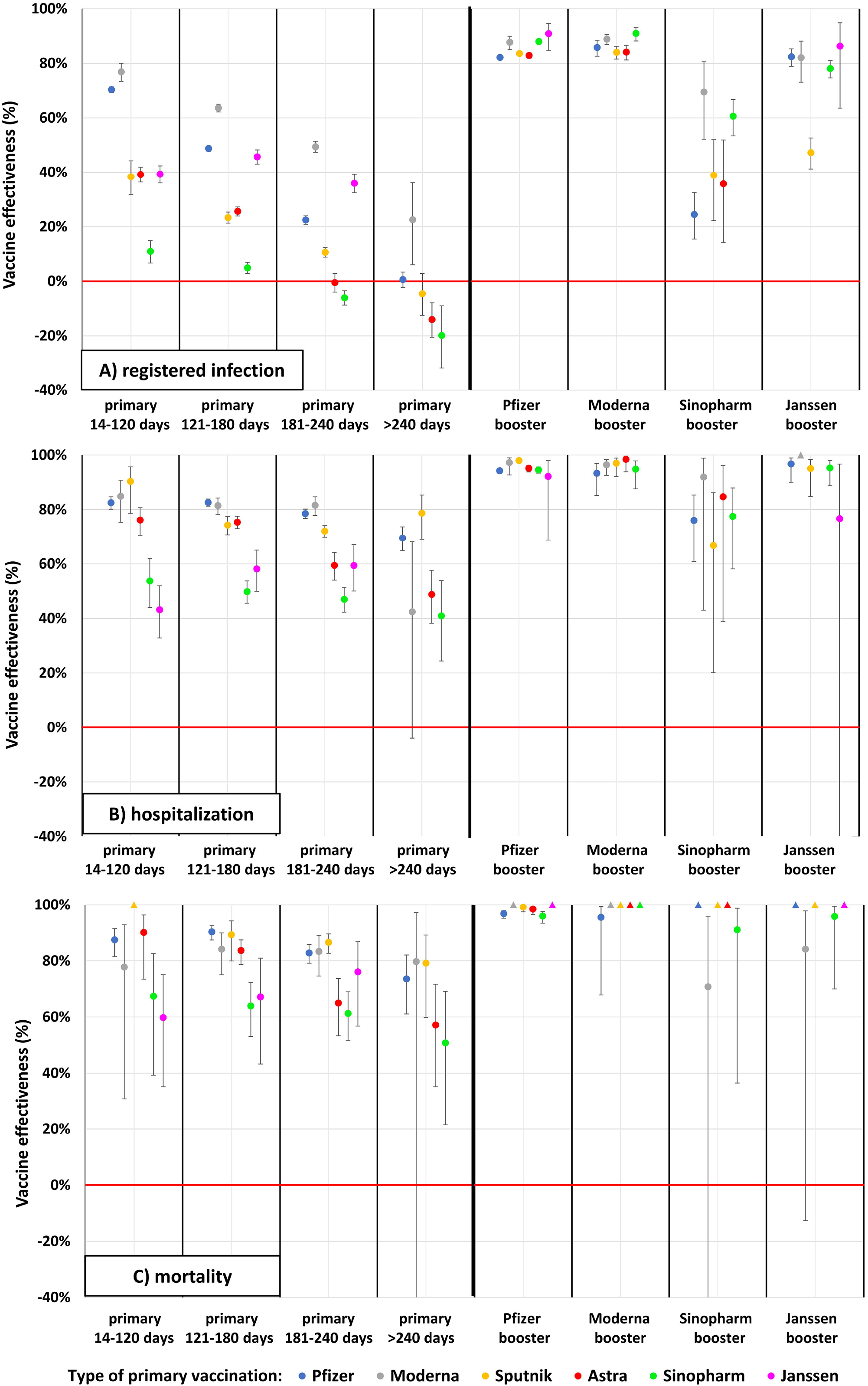
Adjusted vaccine effectiveness against registered SARS-CoV-2 infection, Covid-19 related hospitalization and death during the Delta wave in the Hungarian population aged 16–64 years. *Datapoints marked with triangles at value 100% indicate vaccine combination categories where no outcome occurred. In these categories, the person-time of observation is very limited.

**Figure 2.**
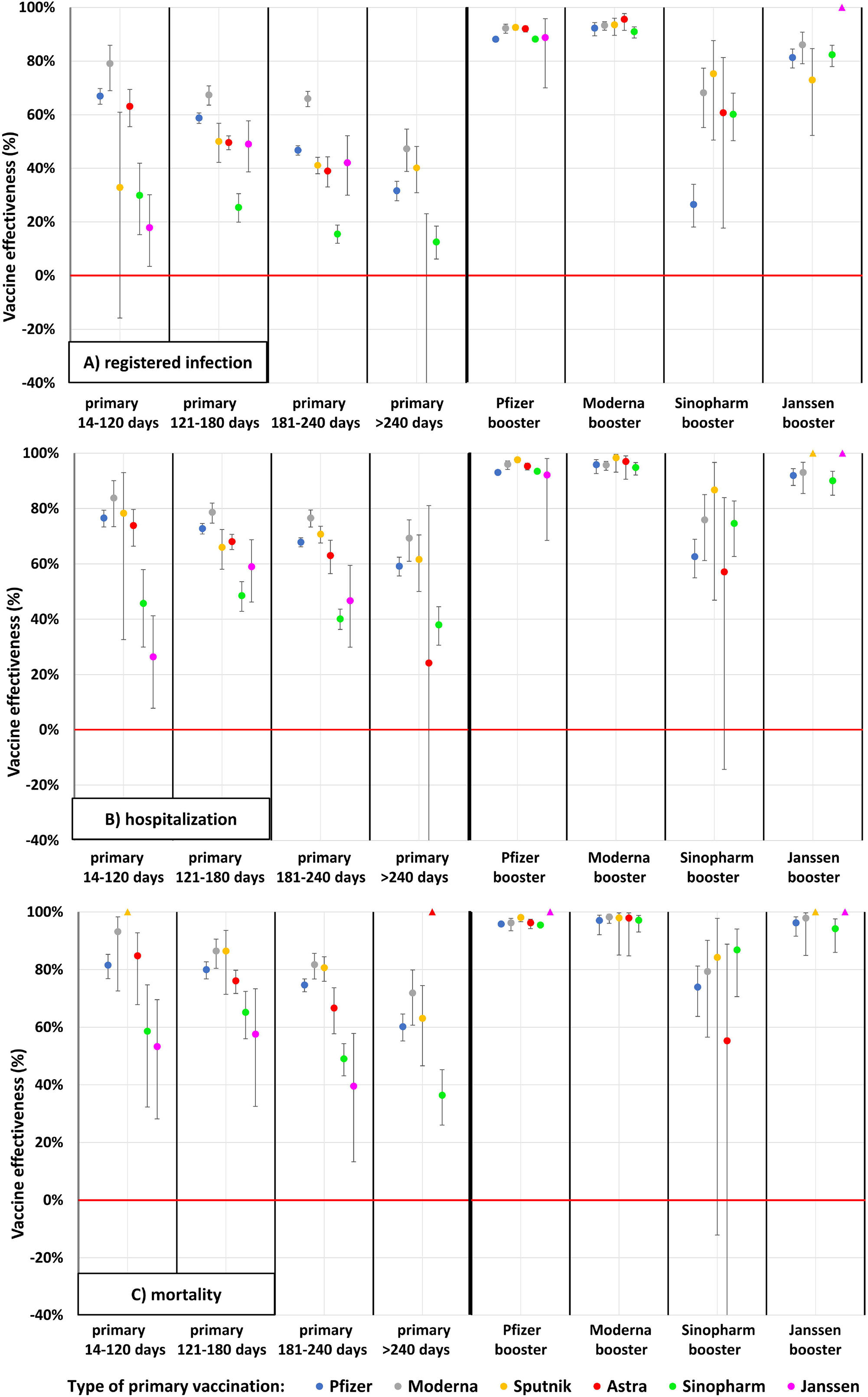
Adjusted vaccine effectiveness against registered SARS-CoV-2 infection, Covid-19 related hospitalization and death during the Delta wave in the Hungarian population aged 65–100 years. *Datapoints marked with triangles at value 100% indicate vaccine combination categories where no outcome occurred. In these categories, the person-time of observation is very limited.

In the primary immunized population aged 65–100 years, adjusted vaccine effectiveness against Covid-19 related hospitalization was 76.6% with the Pfizer-BioNTech, 83.8% with the Moderna, 78.3% with the Sputnik-V, 73.8% with the AstraZeneca, 45.7% with the Sinopharm, and 26.4% with the Janssen vaccine within 14–120 days after the second dose (Figure 2B, Supplementary Table 5). The waning of protection against Covid-19 related hospitalization was much less pronounced than against SARS-CoV-2 infection. Nevertheless, after more than 240 days, primary Pfizer-BioNTech, Moderna, Sputnik-V, and Sinopharm vaccinations had lost around 20% of their effectiveness in relative terms against Covid-19 related hospitalization. Booster vaccination with mRNA vaccines or the Janssen vaccine increased adjusted effectiveness up to almost 100% (92.1% to 97.6% for Pfizer-BioNTech booster, 94.8% to 98.3% for Moderna booster and 90.00% to 100.00% for Janssen booster doses) irrespective of the vaccine type used for primary immunization, while the Sinopharm booster dose proved to be less effective in this regard (57.1% to 86.7%) (Figure 2B, Supplementary Table 5).

In individuals aged 65–100 years, vaccine effectiveness against Covid-19 related death after primary immunization was high or moderate (Pfizer-BioNTech: 81.5%, Moderna: 93.2%, Sputnik-V: 100.0%, AstraZeneca: 84.8%, Sinopharm: 58.6%, Janssen: 53.3%). Vaccine effectiveness decreased to 60.2% for the Pfizer-BioNTech, 71.9% for the Moderna, 63.0% for the Sputnik-V, and 36.4% for the Sinopharm vaccines by >240 days after the completion of primary immunization. All types of booster vaccines restored effectiveness up to almost 100% (95.8% to 100.00% for the Pfizer-BioNTech booster, 97.0% to 98.2% for the Moderna booster and 94.2% to 100.00% for the Janssen booster depending on prior vaccine types), except for the Sinopharm booster (55.3% after 2 doses of AstraZeneca to 86.8% after 2 doses of Sinopharm) (Figure 2C, Supplementary Table 7).

In the age group of 18–64 years, vaccine effectiveness against Covid-19 related death was somewhat higher than in the 65–100 years age group. Similarly to the older population, the Sinopharm and Janssen vaccines showed lower effectiveness compared to other vaccine types. The patterns of waning were very similar in the two age groups, with slightly less waning in terms of Covid-19 related hospitalization and mortality (Figure 1, Supplementary Table 6).

## 4 Discussion

We found lower vaccine effectiveness against the Delta variant (B.1.617.2) within the first 4 months after primary immunization compared to effectiveness rates reported by the HUN-VE 1 study for spring 2021 against the Alpha variant (B.1.1.7) (1), although the results are not directly comparable. The lower effectiveness of globally available vaccine types against the Delta variant compared to Alpha has been widely reported (15,15,17,18,19). Our study is among the first to report effectiveness data for the Sinopharm, Sputnik-V, and Janssen vaccines against the Delta variant, showing a lower benefit for Janssen and Sinopharm compared to other vaccine types.

Most vaccine types provided sustained protection and high effectiveness against Covid-19 related death and hospitalization even after only two doses, which is in line with previous publications (20,21). A recent prospective study from the United States reported similar, 85% effectiveness of mRNA vaccines against Covid-19 related hospitalization among patients infected with the Delta or Alpha SARS-CoV-2 variant after two doses (20). In a study conducted in the United Arab Emirates (UAE), the Sinopharm and Pfizer-BioNTech vaccines provided 95% and 98% effectiveness against Covid-19 related hospitalization in patients infected with the Delta variant, respectively (22). We found high protection against mortality of Pfizer-BioNTech, Moderna, Sputnik-V, and AstraZeneca vaccines but lower than 60% effectiveness for the Janssen and Sinopharm vaccines during the Delta wave in these age groups.

Waning effectiveness was observed for all outcomes, mostly for protection against SARS-CoV-2 infection, especially 6 months after primary immunization. The waning of vaccine effectiveness after primary immunization is widely discussed (23,24), and in most cases it is attributed to the time-dependent decrease of neutralizing response against the SARS-CoV-2 virus. However, most studies have reported maintained and only slightly decreased protection against severe outcomes (4,5,6,25).

There have been very few comparative studies on Sinopharm and other vaccines. The Pfizer vaccine was found to be superior to Sinopharm with respect to post-vaccination quantitative antibody titers (26). Similarly, the Sputnik V vaccine was also more immunogenic compared to Sinopharm (27). Regarding the Omicron variant, both Sinopharm and Janssen yielded significantly lower neutralizing activities compared to Pfizer and Moderna, (28) and, this may explain that in our study, these vaccines showed a steeper decrease in efficacy over time.

Improved protection against all Covid-19 related outcomes with booster vaccines compared to primary immunization has been demonstrated by a number of studies (9,10,11). In a recently published preprint of HUN-VE 2 study from Hungary, booster vaccination provided 96% vaccine effectiveness against SARS-CoV-2 related death as an overall result in comparison to those, having only primary immunization during the Delta wave, where these VE was only 73% (29). Another study from Israel was among the first to report a 93% lower risk of Covid-19 related hospitalization and an 81% lower risk of Covid-19 related death among patients who had received three doses of the Pfizer-BioNTech vaccine compared to those who had received only two doses at least 5 months earlier (30). Later, another Israeli study confirmed the mid-term benefit of booster vaccination with the Pfizer-BioNTech vaccine in the older population (50 years or older), with an adjusted hazard ratio of 0.10 for Covid-19 related death after the third dose vs. primary immunization (11). In a phase 1/2 clinical trial, adults who had received the Pfizer-BioNTech, Janssen, or Moderna vaccine at least 12 weeks prior to enrolment had a 4.2 to 76-fold increase in neutralizing activity against a D614G pseudovirus, and a 4.6 to 56-fold increase in binding antibody titers with all vaccine combinations. Homologous booster doses resulted in a 4.2 to 20-fold increase in neutralizing antibody titers, while heterologous boosters increased titers 6.2 to 76-fold (12). The benefit of booster doses was later confirmed by the VISION Network providing vaccine effectiveness data against Covid-19-associated emergency department and urgent care encounters or hospitalizations during the Delta and Omicron waves (31). In a robust cohort study from the United Kingdom (U.K.), booster vaccination with mRNA vaccines provided 85% to 95% relative effectiveness against symptomatic Covid-19 infection and 97% to 99% absolute effectiveness against hospitalization in all age groups during the Delta wave, irrespective of the primary course, with no sign of waning for up to 10 weeks (32). In line with these observations, our study also demonstrated very high effectiveness against infection with the Delta variant for mRNA booster vaccines during the first 4 months irrespective of the type of the primary immunization, and very high, almost 100% effectiveness against Covid-19 related hospitalization and death. Our results harmonized and partially explained by a recently published Hungarian study, where administering a third dose of BNT162b2 (Pfizer-BioNTech) vaccine following two doses of Sinopharm vaccines provided a significantly enhance both humoral and T cell-mediated immune response, and its effectiveness was comparable to three doses of BNT162b2 vaccines (33).

On the other hand, our study showed less improvement in adjusted vaccine effectiveness after the Sinopharm booster against SARS-CoV-2 infection and Covid-19 related hospitalization and death both in younger and older age groups. Limited data are available on the benefit of the Sinopharm vaccine as a booster option. Two Chinese study demonstrated the safety and high immunogenicity of a third, homologous Sinopharm vaccination among healthy adults (34,35). To our knowledge, this is the first on the effectiveness of the Sinopharm booster against SARS-CoV-2 infection and Covid-19 related hospitalization and death, particularly in comparison with mRNA vaccine boosters.

Although few people received Janssen booster vaccine, we were still able to examine its effectiveness. It proved to be highly effective against SARS-CoV-2 infection as well as against Covid-19 related hospitalization and death, with similar protection to mRNA boosters irrespective of the type of the primary vaccination. Because only few people received AstraZeneca or Sputnik-V boosters in our study, we were not able to assess the effectiveness of these combinations.

The strengths of our study include its nationwide nature inclusion of six different vaccines as primary or booster immunization, the robust number of more than 6 million vaccinated individuals, the 4 different time frames for the evaluation of waning effectiveness after primary vaccination, the 3 different outcomes, and the adjustment for the history of chronic diseases.

However, despite adjustments for age, sex, calendar day, and chronic diseases, further important covariates such as medications or socio-economic status were not included in the analysis. The latter could be related to differences in the likelihood of seeking SARS-CoV-2 testing, chance of detection, uptake of vaccines, prognosis of Covid-19, thus may also have resulted in residual confounding.Furthermore, the diagnosis of SARS-CoV-2 infection could also be established based on clinical symptoms, which might have resulted in differential misclassification, somewhat overestimating vaccine efficacy because physicians might have been less likely to diagnose Covid-19 in vaccinated individuals. On the other hand, misclassification might have biased the results in the other direction (i.e., underestimation of vaccine efficacy), as well, regarding the first infection as outcome, as the proportion of unregistered cases were likely to be higher among the unvaccinated population due to the higher incidence among them. Because of the issue of unregistered cases, we did not censor patients with registered infection in the analysis of the risk of hospitalization and death. As the proportion of individuals with natural immunity were likely to be higher in the unvaccinated population, our estimates can be considered conservative in respect of hospitalization and death. Importantly, vaccine effectiveness was demonstrated when the Delta SARS-CoV-2 variant (B.1.617.2) was the dominant strain in Hungary, therefore, the results do not represent the effectiveness of investigated vaccines against the Omicron variant (B.1.1.529) or against new, upcoming variants.

In conclusion, the nationwide HUN-VE 3 study provides a comprehensive overview of the durability and effectiveness of SARS-CoV-2 vaccines against infection, Covid-19 related hospitalization and mortality with six different vaccine types used for primary immunization and booster doses after primary immunization with various vaccine types. We found significantly waning vaccine effectiveness after primary immunization with each vaccine type, especially against SARS-CoV-2 infection and to a lesser degree against severe Covid-19 related outcomes. On the other hand, our study demonstrates the outstanding benefit of mRNA and Janssen booster vaccines against all Covid-19 related outcomes, irrespective of previously administered vaccine types.

## Data Availability

The datasets generated for this study can be found in the MedRxiv repository. Further inquiries can be directed to the corresponding author.

## 7 Author Contributions

Z Vokó: Conceptualization, Methodology, Formal analysis, Validation and Writing – Review & Editing; Z Kiss and I Wittmann: Conceptualization, Methodology, Investigation, Visualization and Writing – Original Draft; L Polivka Conceptualization, Methodology, Formal analysis, Validation, Data curation; Gy Surján, P Nagy, I Kenessey, A Wéber: Conceptualization, Methodology, Validation, Review & Editing; D Nagy, Gy. Rokszin: Conceptualization, Methodology, Formal analysis; O Surján, V Müller, Z Szekanecz, J Szlávik: Conceptualization, Review & Editing; M Kásler, C Müller: Conceptualization, Methodology, Investigation and Supervision; Zs Barcza: Writing – Original Draft; GA Molnár: Validation, Project Administration

## 9 Acknowledgments

The authors would like to thank Richárd Kiss for the initial data curation and dataset validation.

## 12 Contribution to the field statement

Vaccines became the cornerstone of defense against Covid-19, especially as lockdown measures needed to be eased. In a previous study (HUN-VE 1) we investigated vaccine efficacy of five different vaccines (mRNA-based, subunit-based and inactivated virus-based vaccines) during the alpha wave. In the present paper, we analyze vaccine effectiveness (in terms of infection, hospitalization, and mortality) in primary immunized and first boostered groups of cases during the delta wave using nationwide data from Hungary related to six vaccine types (Pfizer-BioNTech, Moderna, Sputnik-V, AstraZeneca, Sinopharm and in this paper also the Janssen vaccine). We show waning of the protection over time with all vaccines used for primary vaccination. Different combination of primary vaccinations and boosters were effective in a different manner (with Sinopharm and Janssen vaccines somewhat less effective in most aspects). Overall, most boosters restored vaccine effectiveness to nearly 100% in case of Covid-related hospitalization and Covid-related mortality. The strengths of our study are, that it provides nationwide data, it enables comparison of primary vaccinated vs. boostered groups, and that it enables comparison of six different vaccine types. Furthermore, cases with age < 65 or >65 years could be separately analyzed, providing data on vaccine effectiveness in the elderly.

