## Supplementary Tables for "Effectiveness and waning of protection with different SARS-CoV-2 primary and booster vaccines during the Delta pandemic wave in 2021 in Hungary (HUN-VE 3 study)"

**Supplementary Table 1.** Definition of history of chronic diseases included in the analysis

| **Disease** | **ICD codes*** | **Other elements of the definition** |
| --- | --- | --- |
| Acute myocardial infarction | I2100, I2110, I2120, I2130, I2140, I2190, I2191, I2200, I2210, I2280, I2290, I2300, I2310, I2320, I2330, I2340, I2350, I2360, I2380, I2400, I2490 |  |
| Angina | I20 |  |
| Chronic heart failure | I50, I42, I110, J81 |  |
| Peripheral arterial disease | I70 |  |
| Stroke | I61, I6290, I63, I64, I74 |  |
| Asthma bronchiale | J40-44, J47 | the time lag between the first and second occurrences 30-365 days |
| COPD | J45, 46 | the time lag between the first and second occurrences 30-365 days |
| Diabetes mellitus | a) E10-14 or  b) not required  and not polycystic ovary syndrome (E282) or gestational diabetes (O24) | at least 1 prescription of antidiabetic treatment (ATC A10)  at least 2 prescriptions of antidiabetic treatment (ATC A10) |
| Type 1 |  | only insulin or maximum 3 oral antidiabetic prescriptions together with insulin prescriptions in the first half year of treatment |
| Type 2 |  | not Type 1 diabetes |
| Chronic kidney disease | N02, N03, N04, N05, N06, N07, N08, N11, N12, N14, N15, N16, N18, N19, N26, N27, N28, N29 |  |
| Neoplasms | All ICD codes starting with C |  |
| Transplantation | All patients from registry of transplantation |  |
| Immunosuppression | D5900, D5910, D6920, D8690, E05, E0630, G61, G70, I73, K50, K51, K52, K7430, K90, L10, L40, L9590, M0690, M30, M31, M32, M33, M35, M36 |  |

COPD: chronic obstructive pulmonary disease; ICD: International Classification of Diseases; ATC: Anatomical Therapeutic Chemical

* Two occurrences of ICD-10 codes in outpatient or inpatient claims data since 1 January 2013

**Supplementary Table 2.** Incidence, crude and adjusted effectiveness of vaccine combinations against registered SARS-CoV-2 **infection** during the Delta wave in the *18-64 years old* Hungarian population

| **Vaccination** | **Number of cases** | **Average population size (1000 persons)** | **Incidence rate  (per 100 000 person-days)  (95% CI)** | **Crude vaccine efficacy (%) (95% CI)** | **Adjusted vaccine efficacy (%) (95% CI)** |
| --- | --- | --- | --- | --- | --- |
| **Unvaccinated** | 148 198 | 1 992.85 | 67.60 (67.26; 67.95) | reference | reference |
| **Pfizer-BionTech** |  |  |  |  |  |
| **primary 14-120 days** | 3 664 | 289.27 | 11.51 (11.14; 11.89) | 83.0 (82.4; 83.5) | 70.3 (69.2; 71.3) |
| **primary 121-180 days** | 24 917 | 549.89 | 41.19 (40.68; 41.71) | 39.1 (38.2; 39.9) | 48.8 (47.8; 49.7) |
| **primary 181-240 days** | 20 629 | 240.00 | 78.14 (77.08; 79.21) | -15.6 (-17.3; -13.9) | 22.5 (20.9; 24.0) |
| **primary >240 days** | 6 722 | 63.85 | 95.70 (93.43; 98.02) | -41.6 (-45.1; -38.1) | 0.6 (-2.3; 3.4) |
| **Pfizer booster 14-120 days** | 3 240 | 191.11 | 15.41 (14.89; 15.95) | 77.2 (76.4; 78.0) | 82.2 (81.5; 82.8) |
| **Moderna booster 14-120 days** | 91 | 6.65 | 12.45 (10.02; 15.28) | 81.6 (77.4; 85.2) | 85.8 (82.6; 88.4) |
| **Sinopharm booster 14-120 days** | 307 | 4.56 | 61.19 (54.54; 68.43) | 9.5 (-1.2; 19.3) | 24.5 (15.4; 32.5) |
| **Janssen booster 14-120 days** | 118 | 7.08 | 15.16 (12.55; 18.16) | 77.6 (73.1; 81.4) | 82.4 (78.9; 85.3) |
| **Moderna** |  |  |  |  |  |
| **primary 14-120 days** | 191 | 33.48 | 5.19 (4.48; 5.98) | 92.3 (91.2; 93.4) | 76.9 (73.3; 80.0) |
| **primary 121-180 days** | 2 899 | 97.15 | 27.13 (26.15; 28.13) | 59.9 (58.4; 61.3) | 63.6 (62.2; 65.1) |
| **primary 181-240 days** | 2 820 | 46.40 | 55.26 (53.24; 57.33) | 18.3 (15.2; 21.3) | 49.4 (47.3; 51.4) |
| **primary >240 days** | 104 | 1.33 | 71.30 (58.26; 86.39) | -5.5 (-27.8; 13.8) | 22.6 (6.1; 36.2) |
| **Pfizer booster 14-120 days** | 102 | 8.62 | 10.76 (8.77; 13.06) | 84.1 (80.7; 87.0) | 87.7 (85.1; 88.4) |
| **Moderna booster 14-120 days** | 145 | 14.15 | 9.31 (7.86; 10.96) | 86.2 (83.8; 88.4) | 88.9 (86.9; 90.6) |
| **Sinopharm booster 14-120 days** | 19 | 0.69 | 25.15 (15.14; 39.27) | 62.8 (41.9; 77.6) | 69.5 (52.1; 80.5) |
| **Janssen booster 14-120 days** | 23 | 1.35 | 14.06 (8.91; 21.10) | 77.1 (65.7; 85.5) | 82.1 (73.0; 88.1) |
| **Sputnik** |  |  |  |  |  |
| **primary 14-120 days** | 386 | 26.38 | 13.30 (12.01; 14.70) | 80.3 (78.3; 82.2) | 38.3 (31.8; 44.3) |
| **primary 121-180 days** | 8 642 | 275.34 | 28.53 (27.94; 29.14) | 57.8 (56.9; 58.7) | 23.3 (21.3; 25.3) |
| **primary 181-240 days** | 25 412 | 229.77 | 100.54 (99.31; 101.79) | -48.7 (-50.7; -46.8) | 10.6 (8.8; 12.4) |
| **primary >240 days** | 796 | 11.13 | 65.03 (60.59; 69.71) | 3.8 (-3.1; 10.4) | -4.6 (-12.5; 2.9) |
| **Pfizer booster 14-120 days** | 1 826 | 122.90 | 13.51 (12.89; 14.14) | 80.0 (79.1; 80.9) | 83.6 (82.8; 84.4) |
| **Moderna booster 14-120 days** | 184 | 12.42 | 13.46 (11.59; 15.55) | 80.1 (77.0; 82.9) | 84.0 (81.5; 86.2) |
| **Sinopharm booster 14-120 days** | 66 | 1.31 | 45.95 (35.54; 58.46) | 32.0 (13.5; 47.4) | 38.9 (22.2; 52.0) |
| **Janssen booster 14-120 days** | 337 | 6.58 | 46.57 (41.73; 51.82) | 31.1 (23.3; 38.3) | 47.2 (41.2; 52.6) |
| **Astra-Zeneca** |  |  |  |  |  |
| **primary 14-120 days** | 2 205 | 120.53 | 16.63 (15.94; 17.34) | 75.4 (74.3; 76.4) | 39.2 (36.4; 41.9) |
| **primary 121-180 days** | 13 200 | 167.84 | 71.50 (70.28; 72.73) | -5.8 (-7.7; -3.9) | 25.6 (23.9; 27.3) |
| **primary 181-240 days** | 3 995 | 50.16 | 72.40 (70.17; 74.68) | -7.1 (-10.5; -3.8) | -0.5 (-4.0; 2.8) |
| **primary >240 days** | 1 374 | 8.12 | 153.79 (145.76; 162.14) | -127.5 (-139.9; -115.6) | -14.0 (-20.5; -7.9) |
| **Pfizer booster 14-120 days** | 1 372 | 82.96 | 15.03 (14.25; 15.85) | 77.8 (76.5; 78.9) | 82.9 (81.9; 83.8) |
| **Moderna booster 14-120 days** | 141 | 9.11 | 14.08 (11.85; 16.60) | 79.2 (75.4; 82.5) | 84.1 (81.2; 86.5) |
| **Sinopharm booster 14-120 days** | 46 | 0.76 | 55.21 (40.42; 73.64) | 18.3 (-8.9; 40.2) | 35.8 (14.2; 51.9) |
| **Sinopharm** |  |  |  |  |  |
| **primary 14-120 days** | 1 991 | 77.95 | 23.22 (22.21; 24.26) | 65.7 (64.1; 67.2) | 10.9 (6.7; 15.0) |
| **primary 121-180 days** | 17 492 | 220.47 | 72.13 (71.06; 73.20) | -6.7 (-8.4; -5.0) | 4.9 (2.8; 7.0) |
| **primary 181-240 days** | 10 819 | 101.82 | 96.60 (94.79; 98.44) | -42.9 (-45.7; -40.1) | -6.1 (-8.7; -3.4) |
| **primary >240 days** | 448 | 4.24 | 96.03 (87.34; 105.35) | -42.0 (-55.9; -29.2) | -19.9 (-31.9; -9.0) |
| **Pfizer booster 14-120 days** | 1 007 | 98.82 | 9.26 (8.70; 9.85) | 86.3 (85.4; 87.1) | 88.0 (87.2; 88.7) |
| **Moderna booster 14-120 days** | 54 | 6.69 | 7.33 (5.51; 9.57) | 89.2 (85.8; 91.9) | 91.0 (88.2; 93.1) |
| **Sinopharm booster 14-120 days** | 138 | 3.90 | 32.16 (27.01; 37.99) | 52.4 (43.8; 60.0) | 60.6 (53.4; 66.7) |
| **Janssen booster 14-120 days** | 187 | 9.22 | 18.43 (15.88; 21.27) | 72.7 (68.5; 76.5) | 78.1 (74.7; 81.0) |
| **Janssen** |  |  |  |  |  |
| **primary 14-120 days** | 1 571 | 37.27 | 38.32 (36.45; 40.26) | 43.3 (40.4; 46.1) | 39.3 (36.1; 42.4) |
| **primary 121-180 days** | 1 866 | 42.90 | 39.55 (37.77; 41.38) | 41.5 (38.8; 44.1) | 45.7 (43.0; 48.2) |
| **primary 181-240 days** | 1 547 | 21.65 | 64.97 (61.77; 68.29) | 3.9 (-1.0; 8.6) | 35.9 (32.5; 39.2) |
| **Pfizer booster 14-120 days** | 14 | 1.81 | 7.02 (3.84; 11.78) | 89.6 (82.6; 94.3) | 90.9 (84.6; 94.6) |
| **Janssen booster 14-120 days** | 4 | 0.38 | 9.58 (2.61; 24.54) | 85.8 (63.7; 96.1) | 86.3 (63.5; 94.9) |

CI: confidence interval

Vaccine combinations with less than 3,000 persons on 31 December 2021 and less than 300 registered cases since the start of the epidemic were not analyzed.

**Supplementary Table 3.** Incidence, crude and adjusted effectiveness of vaccine combinations against registered SARS-CoV-2 **infection** during the Delta wave in the *65-100 years old* Hungarian population

| **Vaccination** | **Number of cases** | **Average population size (1000 persons)** | **Incidence rate  (per 100 000 person-days) (95% CI)** | **Crude vaccine efficacy (%) (95% CI)** | **Adjusted vaccine efficacy (%) (95% CI)** |
| --- | --- | --- | --- | --- | --- |
| **Unvaccinated** | 15 011 | 249.19 | 54.76 (53.89; 55.65) | reference | reference |
| **Pfizer-BionTech** |  |  |  |  |  |
| **primary 14-120 days** | 518 | 34.54 | 13.63 (12.49; 14.86) | 75.1 (72.8; 77.2) | 67.0 (63.9; 69.8) |
| **primary 121-180 days** | 2 268 | 158.13 | 13.04 (12.51; 13.59) | 76.2 (75.1; 77.2) | 58.8 (56.8; 60.7) |
| **primary 181-240 days** | 6 377 | 144.51 | 40.12 (39.14; 41.11) | 26.7 (24.6; 28.9) | 46.8 (45.0; 48.5) |
| **primary >240 days** | 1 687 | 23.59 | 65.02 (61.96; 68.20) | -18.7 (-24.9; -12.8) | 31.6 (27.9; 35.1) |
| **Pfizer booster 14-120 days** | 2 267 | 220.59 | 9.34 (8.96; 9.74) | 82.9 (82.2; 83.7) | 88.1 (87.6; 88.7) |
| **Moderna booster 14-120 days** | 39 | 5.71 | 6.21 (4.42; 8.49) | 88.7 (84.5; 91.9) | 92.2 (89.4; 94.3) |
| **Sinopharm booster 14-120 days** | 342 | 5.88 | 52.90 (47.44; 58.82) | 3.4 (-7.5; 13.5) | 26.5 (18.1; 34.0) |
| **Janssen booster 14-120 days** | 110 | 7.26 | 13.78 (11.32; 16.61) | 74.8 (69.7; 79.3) | 81.3 (77.4; 84.5) |
| **Moderna** |  |  |  |  |  |
| **primary 14-120 days** | 25 | 5.38 | 4.22 (2.73; 6.23) | 92.3 (88.6; 95.0) | 79.1 (69.0; 85.9) |
| **primary 121-180 days** | 332 | 23.16 | 13.03 (11.67; 14.51) | 76.2 (73.5; 78.7) | 67.4 (63.6; 70.8) |
| **primary 181-240 days** | 604 | 22.40 | 24.52 (22.60; 26.55) | 55.2 (51.4; 58.8) | 66.0 (63.1; 68.7) |
| **primary >240 days** | 175 | 3.14 | 50.62 (43.40; 58.71) | 7.6 (-7.3; 20.8) | 47.3 (38.8; 54.7) |
| **Pfizer booster 14-120 days** | 87 | 12.63 | 6.26 (5.02; 7.72) | 88.6 (85.9; 90.8) | 92.2 (90.4; 93.7) |
| **Moderna booster 14-120 days** | 76 | 12.88 | 5.36 (4.23; 6.71) | 90.2 (87.7; 92.3) | 93.3 (91.5; 94.6) |
| **Sinopharm booster 14-120 days** | 33 | 1.24 | 24.25 (16.69; 34.05) | 55.7 (37.8; 69.5) | 68.2 (55.2; 77.4) |
| **Janssen booster 14-120 days** | 23 | 1.97 | 9.63 (6.10; 14.45) | 80.7 (71.0; 87.7) | 86.1 (79.0; 90.8) |
| **Sputnik** |  |  |  |  |  |
| **primary 14-120 days** | 13 | 0.67 | 17.65 (9.40; 30.18) | 67.8 (44.9; 82.8) | 32.8 (-15.8; 61.0) |
| **primary 121-180 days** | 193 | 28.27 | 6.21 (5.36; 7.15) | 88.7 (86.9; 90.2) | 50.1 (42.3; 56.8) |
| **primary 181-240 days** | 1 817 | 38.08 | 43.37 (41.40; 45.42) | 20.8 (16.8; 24.6) | 41.1 (37.9; 44.2) |
| **primary >240 days** | 194 | 4.35 | 40.56 (35.06; 46.69) | 25.9 (14.7; 36.0) | 40.2 (30.9; 48.2) |
| **Pfizer booster 14-120 days** | 273 | 47.44 | 5.23 (4.63; 5.89) | 90.4 (89.2; 91.6) | 92.5 (91.6; 93.4) |
| **Moderna booster 14-120 days** | 17 | 3.43 | 4.51 (2.63; 7.22) | 91.8 (86.8; 95.2) | 93.5 (89.6; 96.0) |
| **Sinopharm booster 14-120 days** | 8 | 0.44 | 16.58 (7.16; 32.67) | 69.7 (40.3; 86.9) | 75.3 (50.6; 87.7) |
| **Janssen booster 14-120 days** | 12 | 0.58 | 18.83 (9.73; 32.89) | 65.6 (39.9; 82.2) | 72.9 (52.3; 84.6) |
| **Astra-Zeneca** |  |  |  |  |  |
| **primary 14-120 days** | 114 | 23.69 | 4.37 (3.61; 5.26) | 92.0 (90.4; 93.4) | 63.1 (55.6; 69.4) |
| **primary 121-180 days** | 1 811 | 48.49 | 33.95 (32.40; 35.55) | 38.0 (34.9; 41.0) | 49.7 (47.0; 52.2) |
| **primary 181-240 days** | 485 | 8.51 | 51.79 (47.28; 56.61) | 5.4 (-3.5; 13.8) | 38.9 (33.0; 44.3) |
| **primary >240 days** | 7 | 0.04 | 148.46 (59.69; 305.89) | -171.1 (-458.7; -9.0) | -61.5 (-238.9; 23.0) |
| **Pfizer booster 14-120 days** | 194 | 26.85 | 6.57 (5.68; 7.56) | 88.0 (86.2; 89.6) | 92.0 (90.8; 93.1) |
| **Moderna booster 14-120 days** | 9 | 2.24 | 3.65 (1.67; 6.92) | 93.3 (87.4; 97.0) | 95.6 (91.4; 97.7) |
| **Sinopharm booster 14-120 days** | 7 | 0.20 | 31.38 (12.62; 64.65) | 42.7 (-18.1; 77.0) | 60.8 (17.7; 81.3) |
| **Sinopharm** |  |  |  |  |  |
| **primary 14-120 days** | 109 | 6.11 | 16.22 (13.32; 19.57) | 70.4 (64.2; 75.7) | 29.8 (15.2; 41.9) |
| **primary 121-180 days** | 884 | 49.32 | 16.29 (15.24; 17.40) | 70.2 (68.2; 72.2) | 25.4 (19.9; 30.5) |
| **primary 181-240 days** | 3 703 | 65.18 | 51.65 (50.00; 53.34) | 5.7 (2.2; 9.0) | 15.5 (12.0; 18.8) |
| **primary >240 days** | 920 | 11.21 | 74.62 (69.87; 79.60) | -36.3 (-45.6; -27.3) | 12.5 (6.1; 18.4) |
| **Pfizer booster 14-120 days** | 2 037 | 231.33 | 8.01 (7.66; 8.36) | 85.4 (84.7; 86.0) | 88.2 (87.6; 88.7) |
| **Moderna booster 14-120 days** | 75 | 10.78 | 6.33 (4.98; 7.93) | 88.4 (85.5; 90.9) | 90.9 (88.6; 92.8) |
| **Sinopharm booster 14-120 days** | 80 | 2.58 | 28.23 (22.38; 35.13) | 48.5 (35.8; 59.2) | 60.2 (50.4; 68.0) |
| **Janssen booster 14-120 days** | 79 | 6.08 | 11.81 (9.35; 14.72) | 78.4 (73.1; 82.9) | 82.4 (78.0; 85.9) |
| **Janssen** |  |  |  |  |  |
| **primary 14-120 days** | 150 | 3.17 | 43.08 (36.46; 50.55) | 21.3 (7.6; 33.5) | 17.8 (3.4; 30.1) |
| **primary 121-180 days** | 112 | 3.86 | 26.36 (21.71; 31.72) | 51.9 (42.0; 60.4) | 49.1 (38.6; 57.7) |
| **primary 181-240 days** | 107 | 1.92 | 50.55 (41.42; 61.08) | 7.7 (-11.6; 24.4) | 42.1 (29.9; 52.2) |
| **Pfizer booster 14-120 days** | 4 | 0.44 | 8.31 (2.26; 21.28) | 84.8 (61.1; 95.9) | 88.8 (70.0; 95.8) |
| **Janssen booster 14-120 days** | 0 | 0.06 | 0.00 (0.00; 55.01) | 100.0 (-0.5; 100.0) | 100.0 (NA) |

CI: confidence interval

Vaccine combinations with less than 3,000 persons on 31 December 2021 and less than 300 registered cases since the start of the epidemic were not analyzed.

**Supplementary Table 4.** Incidence, crude and adjusted effectiveness of vaccine combinations against COVID-19 related **hospitalization** during the Delta wave in the *18-64 years old* Hungarian population

| **Vaccination** | **Number of cases** | **Average population size (1000 persons)** | **Incidence rate  (per 100 000 person-days) (95% CI)** | **Crude vaccine efficacy (%) (95% CI)** | **Adjusted vaccine efficacy (%) (95% CI)** |
| --- | --- | --- | --- | --- | --- |
| **Unvaccinated** | 12 738 | 2 213.67 | 5.23 (5.14; 5.32) | reference | reference |
| **Pfizer-BionTech** |  |  |  |  |  |
| **primary 14-120 days** | 236 | 360.42 | 0.60 (0.52; 0.68) | 88.6 (87.1; 90.0) | 82.6 (80.1; 84.7) |
| **primary 121-180 days** | 748 | 636.89 | 1.07 (0.99; 1.15) | 79.6 (78.0; 81.1) | 82.6 (81.2; 83.9) |
| **primary 181-240 days** | 671 | 281.14 | 2.17 (2.01; 2.34) | 58.5 (55.2; 61.7) | 78.5 (76.7; 80.2) |
| **primary >240 days** | 194 | 73.72 | 2.39 (2.07; 2.75) | 54.3 (47.3; 60.5) | 69.6 (64.9; 73.6) |
| **Pfizer booster 14-120 days** | 160 | 217.58 | 0.67 (0.57; 0.78) | 87.2 (85.1; 89.1) | 94.3 (93.3; 95.1) |
| **Moderna booster 14-120 days** | 6 | 7.57 | 0.72 (0.26; 1.57) | 86.2 (70.0; 94.9) | 93.3 (85.2; 97.0) |
| **Sinopharm booster 14-120 days** | 16 | 5.36 | 2.71 (1.55; 4.41) | 48.1 (15.7; 70.4) | 76.0 (60.9; 85.3) |
| **Janssen booster 14-120 days** | 3 | 8.06 | 0.34 (0.07; 0.99) | 93.5 (81.1; 98.7) | 96.8 (90.0; 99.0) |
| **Moderna** |  |  |  |  |  |
| **primary 14-120 days** | 16 | 40.74 | 0.36 (0.20; 0.58) | 93.2 (88.9; 96.1) | 84.9 (75.4; 90.8) |
| **primary 121-180 days** | 147 | 110.34 | 1.21 (1.02; 1.42) | 76.8 (72.8; 80.5) | 81.5 (78.2; 84.3) |
| **primary 181-240 days** | 117 | 52.23 | 2.04 (1.68; 2.44) | 61.1 (53.3; 67.8) | 81.6 (77.9; 84.7) |
| **primary >240 days** | 11 | 1.65 | 6.07 (3.03; 10.86) | -16.0 (-107.7; 42.1) | 42.5 (-4.0; 68.2) |
| **Pfizer booster 14-120 days** | 4 | 9.63 | 0.38 (0.10; 0.97) | 92.8 (81.5; 98) | 97.3 (92.7; 99) |
| **Moderna booster 14-120 days** | 7 | 15.92 | 0.40 (0.16; 0.82) | 92.4 (84.2; 96.9) | 96.5 (92.5; 98.3) |
| **Sinopharm booster 14-120 days** | 1 | 0.85 | 1.07 (0.03; 5.95) | 79.6 (-13.7; 99.5) | 92.0 (43.0; 98.9) |
| **Janssen booster 14-120 days** | 0 | 1.52 | 0.00 (0.00; 2.01) | 100.0 (57.8; 100.0) | 100.0 (NA) |
| **Sputnik** |  |  |  |  |  |
| **primary 14-120 days** | 6 | 30.90 | 0.18 (0.06; 0.38) | 96.6 (92.7; 98.8) | 90.4 (78.5; 95.7) |
| **primary 121-180 days** | 241 | 295.38 | 0.74 (0.65; 0.84) | 85.8 (83.9; 87.6) | 74.3 (70.7; 77.5) |
| **primary 181-240 days** | 758 | 253.36 | 2.72 (2.53; 2.92) | 48.0 (44.0; 51.7) | 72.1 (69.9; 74.2) |
| **primary >240 days** | 28 | 13.30 | 1.91 (1.27; 2.77) | 63.4 (47.1; 75.7) | 78.7 (69.1; 85.4) |
| **Pfizer booster 14-120 days** | 30 | 130.59 | 0.21 (0.14; 0.30) | 96.0 (94.3; 97.3) | 98.0 (97.1; 98.6) |
| **Moderna booster 14-120 days** | 4 | 13.27 | 0.27 (0.07; 0.70) | 94.8 (86.6; 98.6) | 97.0 (92.1; 98.9) |
| **Sinopharm booster 14-120 days** | 5 | 1.42 | 3.20 (1.04; 7.46) | 38.9 (-42.6; 80.2) | 66.8 (20.1; 86.2) |
| **Janssen booster 14-120 days** | 3 | 7.23 | 0.38 (0.08; 1.10) | 92.8 (78.9; 98.5) | 95.1 (84.8; 98.4) |
| **Astra-Zeneca** |  |  |  |  |  |
| **primary 14-120 days** | 89 | 128.46 | 0.63 (0.51; 0.78) | 88.0 (85.2; 90.3) | 76.2 (70.6; 80.7) |
| **primary 121-180 days** | 493 | 183.32 | 2.44 (2.23; 2.67) | 53.3 (48.9; 57.4) | 75.4 (73.0; 77.6) |
| **primary 181-240 days** | 255 | 55.78 | 4.16 (3.66; 4.70) | 20.6 (10.1; 30.1) | 59.5 (54.0; 64.2) |
| **primary >240 days** | 111 | 9.55 | 10.57 (8.69; 12.72) | -102.0 (-143.5; -66.0) | 48.8 (38.2; 57.6) |
| **Pfizer booster 14-120 days** | 68 | 88.58 | 0.70 (0.54; 0.88) | 86.7 (83.1; 89.6) | 95.1 (93.8; 96.2) |
| **Moderna booster 14-120 days** | 2 | 9.77 | 0.19 (0.02; 0.67) | 96.4 (87.1; 99.6) | 98.5 (93.9; 99.6) |
| **Sinopharm booster 14-120 days** | 2 | 0.82 | 2.23 (0.27; 8.06) | 57.4 (-54.1; 94.8) | 84.7 (38.9; 96.2) |
| **Sinopharm** |  |  |  |  |  |
| **primary 14-120 days** | 106 | 87.74 | 1.10 (0.90; 1.33) | 79.0 (74.6; 82.8) | 53.8 (43.9; 61.9) |
| **primary 121-180 days** | 655 | 239.02 | 2.49 (2.30; 2.69) | 52.4 (48.5; 56.0) | 49.8 (45.6; 53.8) |
| **primary 181-240 days** | 584 | 114.32 | 4.64 (4.27; 5.04) | 11.2 (3.5; 18.4) | 47.0 (42.3; 51.4) |
| **primary >240 days** | 65 | 4.93 | 12.00 (9.26; 15.29) | -129.4 (-192.5; -76.9) | 40.9 (24.4; 53.8) |
| **Pfizer booster 14-120 days** | 89 | 105.07 | 0.77 (0.62; 0.95) | 85.3 (81.9; 88.2) | 94.6 (93.3; 95.6) |
| **Moderna booster 14-120 days** | 5 | 7.18 | 0.63 (0.21; 1.48) | 87.9 (71.7; 96.1) | 94.8 (87.6; 97.9) |
| **Sinopharm booster 14-120 days** | 10 | 4.23 | 2.15 (1.03; 3.95) | 58.9 (24.5; 80.3) | 77.5 (58.2; 87.9) |
| **Janssen booster 14-120 days** | 5 | 10.01 | 0.45 (0.15; 1.06) | 91.3 (79.7; 97.2) | 95.3 (88.7; 98.0) |
| **Janssen** |  |  |  |  |  |
| **primary 14-120 days** | 140 | 44.57 | 2.86 (2.40; 3.37) | 45.4 (35.5; 54.1) | 43.2 (32.9; 52.0) |
| **primary 121-180 days** | 121 | 49.03 | 2.24 (1.86; 2.68) | 57.1 (48.7; 64.4) | 58.2 (49.9; 65.1) |
| **primary 181-240 days** | 91 | 24.97 | 3.31 (2.67; 4.07) | 36.7 (22.2; 49.1) | 59.4 (50.1; 67.0) |
| **Pfizer booster 14-120 days** | 2 | 2.46 | 0.74 (0.09; 2.67) | 85.9 (49.0; 98.3) | 92.2 (68.8; 98.0) |
| **Janssen booster 14-120 days** | 1 | 0.48 | 1.90 (0.05; 10.61) | 63.6 (-102.9; 99.1) | 76.7 (-65.7; 96.7) |

CI: confidence interval

Vaccine combinations with less than 3,000 persons on 31 December 2021 and less than 300 registered cases since the start of the epidemic were not analyzed.

**Supplementary Table 5.** Incidence, crude and adjusted effectiveness of vaccine combinations against COVID-19 related **hospitalization** during the Delta wave in the *65-100 years old* Hungarian population

| **Vaccination** | **Number of cases** | **Average population size (1000 persons)** | **Incidence rate  (per 100 000 person-days) (95% CI)** | **Crude vaccine efficacy (%) (95% CI)** | **Adjusted vaccine efficacy (%) (95% CI)** |
| --- | --- | --- | --- | --- | --- |
| **Unvaccinated** | 8 269 | 261.16 | 28.78 (28.17; 29.41) | reference | reference |
| **Pfizer-BionTech** |  |  |  |  |  |
| **primary 14-120 days** | 243 | 40.88 | 5.40 (4.75; 6.13) | 81.2 (78.7; 83.5) | 76.6 (73.3; 79.4) |
| **primary 121-180 days** | 949 | 167.53 | 5.15 (4.83; 5.49) | 82.1 (80.9; 83.3) | 72.7 (70.7; 74.6) |
| **primary 181-240 days** | 2 082 | 150.66 | 12.56 (12.03; 13.11) | 56.4 (54.2; 58.4) | 67.8 (66.2; 69.4) |
| **primary >240 days** | 639 | 25.26 | 23.00 (21.25; 24.86) | 20.1 (13.4; 26.4) | 59.2 (55.6; 62.4) |
| **Pfizer booster 14-120 days** | 794 | 232.40 | 3.11 (2.89; 3.33) | 89.2 (88.4; 90.0) | 93.0 (92.5; 93.5) |
| **Moderna booster 14-120 days** | 12 | 5.95 | 1.83 (0.95; 3.20) | 93.6 (88.9; 96.7) | 95.8 (92.6; 97.6) |
| **Sinopharm booster 14-120 days** | 115 | 6.51 | 16.06 (13.26; 19.27) | 44.2 (32.9; 54.0) | 62.6 (55.0; 68.9) |
| **Janssen booster 14-120 days** | 29 | 7.66 | 3.44 (2.30; 4.94) | 88,0 (82.8; 92.0) | 91.9 (88.3; 94.4) |
| **Moderna** |  |  |  |  |  |
| **primary 14-120 days** | 16 | 6.66 | 2.18 (1.25; 3.55) | 92.4 (87.7; 95.7) | 83.8 (73.5; 90.1) |
| **primary 121-180 days** | 139 | 25.45 | 4.96 (4.17; 5.86) | 82.8 (79.6; 85.5) | 78.6 (74.7; 82.0) |
| **primary 181-240 days** | 229 | 23.33 | 8.92 (7.81; 10.16) | 69.0 (64.6; 72.9) | 76.6 (73.3; 79.5) |
| **primary >240 days** | 67 | 3.31 | 18.42 (14.28; 23.40) | 36.0 (18.6; 50.4) | 69.3 (60.9; 75.9) |
| **Pfizer booster 14-120 days** | 28 | 13.33 | 1.91 (1.27; 2.76) | 93.4 (90.4; 95.6) | 96.0 (94.1; 97.2) |
| **Moderna booster 14-120 days** | 29 | 13.69 | 1.93 (1.29; 2.77) | 93.3 (90.4; 95.5) | 95.7 (93.8; 97) |
| **Sinopharm booster 14-120 days** | 17 | 1.39 | 11.15 (6.50; 17.86) | 61.3 (37.9; 77.4) | 75.9 (61.2; 85.0) |
| **Janssen booster 14-120 days** | 7 | 2.06 | 2.81 (1.13; 5.79) | 89.3 (77.8; 95.7) | 93.0 (85.4; 96.7) |
| **Sputnik** |  |  |  |  |  |
| **primary 14-120 days** | 3 | 0.89 | 3.06 (0.63; 8.93) | 89.4 (69.0; 97.8) | 78.3 (32.7; 93.0) |
| **primary 121-180 days** | 92 | 29.25 | 2.86 (2.30; 3.51) | 90.1 (87.8; 92.0) | 66.0 (58.1; 72.4) |
| **primary 181-240 days** | 402 | 39.25 | 9.31 (8.42; 10.27) | 67.7 (64.2; 70.8) | 70.7 (67.5; 73.6) |
| **primary >240 days** | 57 | 4.68 | 11.08 (8.39; 14.35) | 61.5 (50.1; 70.9) | 61.6 (50.0; 70.5) |
| **Pfizer booster 14-120 days** | 40 | 48.72 | 0.75 (0.53; 1.02) | 97.4 (96.5; 98.1) | 97.6 (96.7; 98.2) |
| **Moderna booster 14-120 days** | 2 | 3.52 | 0.52 (0.06; 1.86) | 98.2 (93.5; 99.8) | 98.3 (93.1; 99.6) |
| **Sinopharm booster 14-120 days** | 2 | 0.46 | 3.95 (0.48; 14.25) | 86.3 (50.5; 98.3) | 86.7 (46.9; 96.7) |
| **Janssen booster 14-120 days** | 0 | 0.61 | 0.00 (0.00; 5.51) | 100.0 (80.9; 100.0) | 100.0 (NA) |
| **Astra-Zeneca** |  |  |  |  |  |
| **primary 14-120 days** | 63 | 24.54 | 2.33 (1.79; 2.99) | 91.9 (89.6; 93.8) | 73.8 (66.4; 79.7) |
| **primary 121-180 days** | 588 | 50.15 | 10.66 (9.81; 11.56) | 63.0 (59.7; 66.0) | 68.1 (65.2; 70.7) |
| **primary 181-240 days** | 152 | 9.02 | 15.32 (12.98; 17.96) | 46.8 (37.5; 55.0) | 63.0 (56.5; 68.6) |
| **primary >240 days** | 2 | 0.05 | 39.67 (4.80; 143.32) | -37.8 (-398.1; 83.3) | 24.2 (-203.1; 81.1) |
| **Pfizer booster 14-120 days** | 57 | 27.75 | 1.87 (1.41; 2.42) | 93.5 (91.6; 95.1) | 95.3 (93.9; 96.4) |
| **Moderna booster 14-120 days** | 3 | 2.31 | 1.18 (0.24; 3.45) | 95.9 (88.0; 99.2) | 97.0 (90.6; 99) |
| **Sinopharm booster 14-120 days** | 4 | 0.21 | 17.18 (4.68; 43.99) | 40.3 (-52.9; 83.7) | 57.1 (-14.4; 83.9) |
| **Sinopharm** |  |  |  |  |  |
| **primary 14-120 days** | 60 | 7.14 | 7.64 (5.83; 9.84) | 73.4 (65.8; 79.8) | 45.7 (30.0; 57.9) |
| **primary 121-180 days** | 393 | 52.06 | 6.86 (6.20; 7.58) | 76.2 (73.6; 78.5) | 48.5 (42.8; 53.6) |
| **primary 181-240 days** | 1 358 | 67.36 | 18.33 (17.37; 19.33) | 36.3 (32.6; 39.9) | 40.1 (36.4; 43.6) |
| **primary >240 days** | 341 | 12.07 | 25.69 (23.04; 28.57) | 10.7 (0.5; 20.1) | 38.0 (30.6; 44.5) |
| **Pfizer booster 14-120 days** | 579 | 237.05 | 2.22 (2.04; 2.41) | 92.3 (91.6; 92.9) | 93.4 (92.8; 94.0) |
| **Moderna booster 14-120 days** | 22 | 11.07 | 1.81 (1.13; 2.74) | 93.7 (90.5; 96.1) | 94.8 (92.1; 96.6) |
| **Sinopharm booster 14-120 days** | 26 | 2.68 | 8.83 (5.77; 12.94) | 69.3 (55; 80.0) | 74.6 (62.7; 82.7) |
| **Janssen booster 14-120 days** | 22 | 6.27 | 3.19 (2.00; 4.83) | 88.9 (83.2; 93.1) | 90.0 (84.8; 93.4) |
| **Janssen** |  |  |  |  |  |
| **primary 14-120 days** | 77 | 3.53 | 19.83 (15.65; 24.78) | 31.1 (13.8; 45.7) | 26.4 (7.9; 41.3) |
| **primary 121-180 days** | 53 | 4.44 | 10.86 (8.14; 14.21) | 62.3 (50.6; 71.8) | 59.0 (46.2; 68.7) |
| **primary 181-240 days** | 52 | 2.17 | 21.81 (16.29; 28.6) | 24.2 (0.5; 43.5) | 46.7 (29.9; 59.5) |
| **Pfizer booster 14-120 days** | 2 | 0.62 | 2.92 (0.35; 10.56) | 89.8 (63.3; 98.8) | 92.1 (68.5; 98.0) |
| **Janssen booster 14-120 days** | 0 | 0.07 | 0 (0.00; 45.92) | 100.0 (-59.6; 100.0) | 100.0 (NA) |

CI: confidence interval

Vaccine combinations with less than 3,000 persons on 31 December 2021 and less than 300 registered cases since the start of the epidemic were not analyzed.

**Supplementary Table 6.** Incidence, crude and adjusted effectiveness of vaccine combinations against COVID-19 related **mortality** during the Delta wave in the *18-64 years old* Hungarian population

| **Vaccination** | **Number of cases** | **Average population size (1000 persons)** | **Incidence rate  (per 100 000 person-days) (95% CI)** | **Crude vaccine efficacy (%) (95% CI)** | **Adjusted vaccine efficacy (%) (95% CI)** |
| --- | --- | --- | --- | --- | --- |
| **Unvaccinated** | 1 928 | 2 228.48 | 0.79 (0.75; 0.82) | reference | reference |
| **Pfizer-BionTech** |  |  |  |  |  |
| **primary 14-120 days** | 26 | 366.56 | 0.06 (0.04; 0.09) | 91.8 (87.9; 94.7) | 87.4 (81.5; 91.5) |
| **primary 121-180 days** | 59 | 640.70 | 0.08 (0.06; 0.11) | 89.4 (86.2; 91.9) | 90.3 (87.4; 92.5) |
| **primary 181-240 days** | 111 | 281.95 | 0.36 (0.29; 0.43) | 54.5 (44.9; 62.8) | 82.8 (79.1; 85.8) |
| **primary >240 days** | 26 | 73.90 | 0.32 (0.21; 0.47) | 59.3 (40.2; 73.5) | 73.6 (61.1; 82.1) |
| **Pfizer booster 14-120 days** | 23 | 218.63 | 0.10 (0.06; 0.14) | 87.8 (81.7; 92.3) | 96.8 (95.2; 97.9) |
| **Moderna booster 14-120 days** | 1 | 7.61 | 0.12 (0.00; 0.67) | 84.8 (15.3; 99.6) | 95.5 (67.9; 99.4) |
| **Sinopharm booster 14-120 days** | 0 | 5.38 | 0.00 (0.00; 0.62) | 100.0 (20.7; 100.0) | 100.0 (NA) |
| **Janssen booster 14-120 days** | 0 | 8.09 | 0.00 (0.00; 0.41) | 100.0 (47.2; 100.0) | 100.0 (NA) |
| **Moderna** |  |  |  |  |  |
| **primary 14-120 days** | 3 | 41.33 | 0.07 (0.01; 0.19) | 91.6 (75.4; 98.3) | 77.7 (30.7; 92.8) |
| **primary 121-180 days** | 19 | 111.11 | 0.16 (0.09; 0.24) | 80.2 (69.0; 88.1) | 84.1 (75.0; 89.9) |
| **primary 181-240 days** | 22 | 52.40 | 0.38 (0.24; 0.58) | 51.5 (26.3; 69.7) | 83.3 (74.6; 89.0) |
| **primary >240 days** | 1 | 1.66 | 0.55 (0.01; 3.06) | 30.2 (-289.4; 98.2) | 79.7 (-44.3; 97.1) |
| **Pfizer booster 14-120 days** | 0 | 9.72 | 0.00 (0.00; 0.35) | 100.0 (56.1; 100.0) | 100.0 (NA) |
| **Moderna booster 14-120 days** | 0 | 16.03 | 0.00 (0.00; 0.21) | 100.0 (73.4; 100.0) | 100.0 (NA) |
| **Sinopharm booster 14-120 days** | 1 | 0.86 | 1.05 (0.03; 5.86) | -33.6 (-645.4; 96.6) | 70.7 (-107.9; 95.9) |
| **Janssen booster 14-120 days** | 1 | 1.53 | 0.54 (0.01; 3.01) | 24.5 (-321.4; 98.1) | 84.1 (-12.6; 97.8) |
| **Sputnik** |  |  |  |  |  |
| **primary 14-120 days** | 0 | 31.19 | 0.00 (0.00; 0.11) | 100.0 (86.3; 100.0) | 100.0 (NA) |
| **primary 121-180 days** | 10 | 295.81 | 0.03 (0.01; 0.06) | 96.1 (92.8; 98.1) | 89.3 (79.9; 94.3) |
| **primary 181-240 days** | 62 | 253.79 | 0.22 (0.17; 0.28) | 71.8 (63.6; 78.4) | 86.5 (82.7; 89.6) |
| **primary >240 days** | 9 | 13.37 | 0.61 (0.28; 1.16) | 22.2 (-48.0; 64.5) | 79.1 (59.8; 89.2) |
| **Pfizer booster 14-120 days** | 3 | 130.74 | 0.02 (0.00; 0.06) | 97.3 (92.2; 99.5) | 99.2 (97.4; 99.7) |
| **Moderna booster 14-120 days** | 0 | 13.28 | 0.00 (0.00; 0.25) | 100.0 (67.9; 100.0) | 100.0 (NA) |
| **Sinopharm booster 14-120 days** | 0 | 1.43 | 0.00 (0.00; 2.35) | 100.0 (-199.1; 100.0) | 100.0 (NA) |
| **Janssen booster 14-120 days** | 0 | 7.24 | 0.00 (0.00; 0.46) | 100.0 (41.0; 100.0) | 100.0 (NA) |
| **Astra-Zeneca** |  |  |  |  |  |
| **primary 14-120 days** | 4 | 128.71 | 0.03 (0.01; 0.07) | 96.4 (90.8; 99.0) | 90.1 (73.5; 96.3) |
| **primary 121-180 days** | 57 | 183.79 | 0.28 (0.21; 0.37) | 64.2 (53.4; 73.0) | 83.6 (78.7; 87.4) |
| **primary 181-240 days** | 48 | 56.01 | 0.78 (0.57; 1.03) | 0.9 (-31.9; 27.2) | 65.0 (53.3; 73.7) |
| **primary >240 days** | 23 | 9.64 | 2.17 (1.38; 3.26) | -175.9 (-315.2; -74.5) | 57.1 (35.2; 71.6) |
| **Pfizer booster 14-120 days** | 6 | 88.79 | 0.06 (0.02; 0.13) | 92.2 (83.0; 97.1) | 98.4 (96.5; 99.3) |
| **Moderna booster 14-120 days** | 0 | 9.79 | 0.00 (0.00; 0.34) | 100.0 (56.4; 100.0) | 100.0 (NA) |
| **Sinopharm booster 14-120 days** | 0 | 0.82 | 0.00 (0.00; 4.11) | 100.0 (-422.8; 100.0) | 100.0 (NA) |
| **Sinopharm** |  |  |  |  |  |
| **primary 14-120 days** | 10 | 88.32 | 0.10 (0.05; 0.19) | 86.9 (75.9; 93.7) | 67.4 (39.2; 82.5) |
| **primary 121-180 days** | 57 | 239.80 | 0.22 (0.16; 0.28) | 72.5 (64.2; 79.3) | 63.9 (53.0; 72.3) |
| **primary 181-240 days** | 81 | 114.78 | 0.64 (0.51; 0.80) | 18.4 (-1.9; 35.5) | 61.2 (51.6; 69.0) |
| **primary >240 days** | 18 | 5.01 | 3.27 (1.94; 5.17) | -315.6 (-558.7; -145.8) | 50.7 (21.4; 69.1) |
| **Pfizer booster 14-120 days** | 17 | 105.50 | 0.15 (0.09; 0.23) | 81.4 (70.1; 89.2) | 95.9 (93.4; 97.5) |
| **Moderna booster 14-120 days** | 0 | 7.21 | 0.00 (0.00; 0.46) | 100.0 (40.8; 100.0) | 100.0 (NA) |
| **Sinopharm booster 14-120 days** | 1 | 4.27 | 0.21 (0.01; 1.19) | 72.9 (-51.1; 99.3) | 91.1 (36.4; 98.7) |
| **Janssen booster 14-120 days** | 1 | 10.03 | 0.09 (0.00; 0.50) | 88.5 (35.7; 99.7) | 95.8 (70.0; 99.4) |
| **Janssen** |  |  |  |  |  |
| **primary 14-120 days** | 17 | 45.12 | 0.34 (0.20; 0.55) | 56.5 (30.1; 74.7) | 59.8 (35.2; 75.1) |
| **primary 121-180 days** | 13 | 49.41 | 0.24 (0.13; 0.41) | 69.6 (47.9; 83.8) | 67.1 (43.2; 81.0) |
| **primary 181-240 days** | 11 | 25.14 | 0.40 (0.20; 0.71) | 49.4 (9.3; 74.8) | 76.1 (56.7; 86.8) |
| **Pfizer booster 14-120 days** | 0 | 2.53 | 0.00 (0.00; 1.33) | 100.0 (-68.6; 100.0) | 100.0 (NA) |
| **Janssen booster 14-120 days** | 0 | 0.48 | 0.00 (0.00; 6.93) | 100.0 (-782.6; 100.0) | 100.0 (NA) |

CI: confidence interval

Vaccine combinations with less than 3,000 persons on 31 December 2021 and less than 300 registered cases since the start of the epidemic were not analyzed.

**Supplementary Table 7.** Incidence, crude and adjusted effectiveness of vaccine combinations against COVID-19 related **mortality** during the Delta wave in the *65-100 years old* Hungarian population

| **Vaccination** | **Number of cases** | **Average population size (1000 persons)** | **Incidence rate  (per 100 000 person-days) (95% CI)** | **Crude vaccine efficacy (%) (95% CI)** | **Adjusted vaccine efficacy (%) (95% CI)** |
| --- | --- | --- | --- | --- | --- |
| **Unvaccinated** | 3 366 | 266.84 | 11.47 (11.08; 11.86) | reference | reference |
| **Pfizer-BionTech** |  |  |  |  |  |
| **primary 14-120 days** | 77 | 43.96 | 1.59 (1.26; 1.99) | 86.1 (82.6; 89.1) | 81.5 (76.8; 85.3) |
| **primary 121-180 days** | 195 | 169.79 | 1.04 (0.90; 1.20) | 90.9 (89.5; 92.2) | 79.9 (76.8; 82.7) |
| **primary 181-240 days** | 625 | 151.89 | 3.74 (3.45; 4.05) | 67.4 (64.5; 70.1) | 74.6 (72.3; 76.8) |
| **primary >240 days** | 321 | 25.73 | 11.34 (10.13; 12.65) | 1.1 (-10.9; 12.1) | 60.2 (55.2; 64.6) |
| **Pfizer booster 14-120 days** | 226 | 234.09 | 0.88 (0.77; 1.00) | 92.3 (91.2; 93.3) | 95.8 (95.2; 96.3) |
| **Moderna booster 14-120 days** | 4 | 5.98 | 0.61 (0.17; 1.56) | 94.7 (86.4; 98.6) | 97.0 (92.1; 98.9) |
| **Sinopharm booster 14-120 days** | 36 | 6.57 | 4.98 (3.49; 6.89) | 56.6 (39.8; 69.6) | 73.9 (63.8; 81.2) |
| **Janssen booster 14-120 days** | 6 | 7.71 | 0.71 (0.26; 1.54) | 93.8 (86.6; 97.7) | 96.2 (91.6; 98.3) |
| **Moderna** |  |  |  |  |  |
| **primary 14-120 days** | 2 | 7.15 | 0.25 (0.03; 0.92) | 97.8 (92.0; 99.7) | 93.2 (72.6; 98.3) |
| **primary 121-180 days** | 29 | 26.14 | 1.01 (0.68; 1.45) | 91.2 (87.3; 94.1) | 86.4 (80.4; 90.6) |
| **primary 181-240 days** | 68 | 23.54 | 2.63 (2.04; 3.33) | 77.1 (70.9; 82.3) | 81.7 (76.7; 85.6) |
| **primary >240 days** | 35 | 3.37 | 9.45 (6.58; 13.15) | 17.6 (-14.9; 42.7) | 71.9 (60.7; 79.9) |
| **Pfizer booster 14-120 days** | 13 | 13.49 | 0.88 (0.47; 1.50) | 92.4 (86.9; 95.9) | 96.2 (93.5; 97.8) |
| **Moderna booster 14-120 days** | 6 | 13.87 | 0.39 (0.14; 0.86) | 96.6 (92.5; 98.7) | 98.2 (96.1; 99.2) |
| **Sinopharm booster 14-120 days** | 7 | 1.41 | 4.51 (1.81; 9.28) | 60.7 (18.9; 84.2) | 79.3 (56.6; 90.1) |
| **Janssen booster 14-120 days** | 1 | 2.08 | 0.40 (0.01; 2.22) | 96.2 (78.7; 99.9) | 97.9 (84.9; 99.7) |
| **Sputnik** |  |  |  |  |  |
| **primary 14-120 days** | 0 | 0.98 | 0.00 (0.00; 3.41) | 100.0 (70.2; 100.0) | 100.0 (NA) |
| **primary 121-180 days** | 7 | 29.40 | 0.22 (0.09; 0.45) | 98.1 (96.1; 99.2) | 86.4 (71.4; 93.6) |
| **primary 181-240 days** | 86 | 39.43 | 1.98 (1.59; 2.45) | 82.7 (78.6; 86.2) | 80.7 (76.0; 84.4) |
| **primary >240 days** | 29 | 4.75 | 5.55 (3.72; 7.98) | 51.6 (30.3; 67.6) | 63.0 (46.6; 74.4) |
| **Pfizer booster 14-120 days** | 13 | 48.85 | 0.24 (0.13; 0.41) | 97.9 (96.4; 98.9) | 98.1 (96.6; 98.9) |
| **Moderna booster 14-120 days** | 1 | 3.54 | 0.26 (0.01; 1.43) | 97.8 (87.5; 99.9) | 97.9 (85.1; 99.7) |
| **Sinopharm booster 14-120 days** | 1 | 0.46 | 1.97 (0.05; 10.96) | 82.8 (4.3; 99.6) | 84.2 (-12.1; 97.8) |
| **Janssen booster 14-120 days** | 0 | 0.61 | 0.00 (0.00; 5.49) | 100.0 (52.1; 100.0) | 100.0 (NA) |
| **Astra-Zeneca** |  |  |  |  |  |
| **primary 14-120 days** | 7 | 24.70 | 0.26 (0.1; 0.53) | 97.8 (95.4; 99.1) | 84.8 (67.8; 92.8) |
| **primary 121-180 days** | 144 | 50.48 | 2.59 (2.19; 3.05) | 77.4 (73.3; 81.0) | 76.1 (71.7; 79.8) |
| **primary 181-240 days** | 71 | 9.17 | 7.04 (5.5; 8.88) | 38.6 (22.3; 52.2) | 66.7 (57.8; 73.7) |
| **primary >240 days** | 0 | 0.05 | 0.00 (0.00; 71.37) | 100.0 (-522.7; 100.0) | 100.0 (NA) |
| **Pfizer booster 14-120 days** | 22 | 27.87 | 0.72 (0.45; 1.09) | 93.7 (90.5; 96.1) | 96.2 (94.2; 97.5) |
| **Moderna booster 14-120 days** | 1 | 2.31 | 0.39 (0.01; 2.19) | 96.6 (80.9; 99.9) | 97.9 (84.8; 99.7) |
| **Sinopharm booster 14-120 days** | 2 | 0.21 | 8.50 (1.03; 30.70) | 25.9 (-167.9; 91) | 55.3 (-78.9; 88.8) |
| **Sinopharm** |  |  |  |  |  |
| **primary 14-120 days** | 16 | 7.68 | 1.90 (1.08; 3.08) | 83.5 (73.1; 90.6) | 58.6 (32.3; 74.7) |
| **primary 121-180 days** | 74 | 52.92 | 1.27 (1.00; 1.6) | 88.9 (86.0; 91.3) | 65.2 (56.0; 72.4) |
| **primary 181-240 days** | 379 | 68.04 | 5.06 (4.57; 5.6) | 55.8 (50.9; 60.4) | 49.0 (43.2; 54.3) |
| **primary >240 days** | 187 | 12.40 | 13.71 (11.82; 15.83) | -19.6 (-38.6; -2.7) | 36.4 (26.0; 45.3) |
| **Pfizer booster 14-120 days** | 158 | 238.30 | 0.60 (0.51; 0.70) | 94.7 (93.8; 95.5) | 95.4 (94.6; 96.1) |
| **Moderna booster 14-120 days** | 5 | 11.14 | 0.41 (0.13; 0.95) | 96.4 (91.7; 98.8) | 97.1 (93.1; 98.8) |
| **Sinopharm booster 14-120 days** | 6 | 2.70 | 2.02 (0.74; 4.40) | 82.4 (61.6; 93.5) | 86.8 (70.6; 94.1) |
| **Janssen booster 14-120 days** | 5 | 6.30 | 0.72 (0.23; 1.68) | 93.7 (85.3; 98.0) | 94.2 (86.0; 97.6) |
| **Janssen** |  |  |  |  |  |
| **primary 14-120 days** | 21 | 3.73 | 5.12 (3.17; 7.83) | 55.3 (31.6; 72.4) | 53.3 (28.2; 69.6) |
| **primary 121-180 days** | 18 | 4.64 | 3.53 (2.09; 5.57) | 69.2 (51.3; 81.8) | 57.6 (32.5; 73.3) |
| **primary 181-240 days** | 30 | 2.25 | 12.13 (8.18; 17.31) | -5.7 (-51.2; 28.8) | 39.5 (13.3; 57.8) |
| **Pfizer booster 14-120 days** | 0 | 0.69 | 0.00 (0.00; 4.89) | 100.0 (57.4; 100.0) | 100.0 (NA) |
| **Janssen booster 14-120 days** | 0 | 0.08 | 0.00 (0.00; 42.2) | 100.0 (-268.2; 100.0) | 100.0 (NA) |

CI: confidence interval

Vaccine combinations with less than 3,000 persons on 31 December 2021 and less than 300 registered cases since the start of the epidemic were not analyzed.
